# Use of the Spatial Access Ratio to Measure Geospatial Access to Emergency Surgical Services in California

**DOI:** 10.1101/2020.05.29.20116970

**Authors:** Neng Wan, Steven Lizotte, Jiuying Han, Thomas Varghese, Raminder Nirula, Marta McCrum

**Author notes:** Corresponding Author: **Marta McCrum, MD MPH^1^** 30N 1900E Salt Lake City UT 84132.

## Abstract

**Background:** Emergency general surgery (EGS) diseases carry a substantial public health burden, accounting for over 3 million admissions annually. Due to their time-sensitive nature, ensuring adequate access to EGS services is critical for reducing patient morbidity and mortality. Travel-time alone, without consideration of resource supply and demand, may be insufficient to determine a regional health care system’s ability to provide timely access to EGS care. Spatial Access Ratio (SPAR) incorporates travel-time, as well as hospital-specific resources and capacity, to determine healthcare accessibility which may be more appropriate for surgical specialties. We therefore compared SPAR to travel-time in their ability to differentiate spatial access to EGS care for vulnerable populations.

**Methods:** We constructed a Geographic Information Science (GIS) platform using existing road networks, and mapped population location, race and socioeconomic characteristics, as well as all EGS-capable hospitals in California. We then compared the shortest travel time method to the gravity-based SPAR in their ability to identify disparities in spatial access for the population as a whole, and subsequently to describe socio-demographic disparities. Reduced spatial access was defined at > 60 minutes travel time, or lowest three classes of SPAR.

**Results:** 283 EGS-capable hospitals were mapped, 142 (50%) of which had advanced resources. Using shortest travel time, 36.98M people (94.8%) were within 20-minutes driving time to any EGS capable hospital, and 33.49M (85.9%) to an advanced-resourced center. Only 166, 950 (0.4%) experienced prolonged (>60 minutes) travel time to any EGS-capable hospital, which increased to 1.05M (2.7%) for advanced-resources. Using SPAR, 11.5M (29.5%) of people had reduced spatial access to any EGS hospital, which increased to 13.9M (35.7%) when evaluating advanced-resource hospitals. The greatest disparities in spatial access to care were found for rural residents and Native Americans for both overall and advanced EGS services.

**Conclusions:** While travel time and SPAR showed similar overall patterns of spatial access to EGS-capable hospitals, SPAR showed greater differentiation of spatial access across the state. Nearly one-third of California residents have limited or poor access to EGS hospitals, with the greatest disparities noted for Native American and rural residents. These findings argue for the use of gravity-based models such as SPAR that incorporate measures of population demand and hospital capacity when assessing spatial access to surgical services, and have implications for the allocation of healthcare resources to address disparities.

## Introduction

Emergency general surgery (EGS) conditions constitute a significant public health burden in the United States, accounting for 3 million admissions annually with disproportionately higher morbidity and mortality compared to elective procedures.^1–3^ While EGS conditions span a wide range of acuity, data suggests that up to 44% of admissions are high-acuity and pose an imminent threat to life if emergent surgical intervention is not achieved.^4,5^ Timely access to surgical care is therefore critical to optimizing outcomes for this high-risk population.

Unfortunately, hospitals with surgical capabilities are unequally distributed across the United States, leading to considerable disparities in access to care. Limitations in county-level availability of EGS services has been shown to extend beyond rurality and also affect minority and low socioeconomic status populations.^6^ As these same populations are also at risk for worse clinical outcomes, including higher EGS morbidity and mortality, there is growing concern that reduced spatial access to care may be an important driver for worsened outcomes. Developing methods to accurately and meaningfully quantify spatial access to surgical care is critical to evaluating the influence of spatial access on clinical outcomes, yet our understanding of spatial access to surgical services has been largely based on rudimentary measures.

Geographic Information Science (GIS) offers unique opportunities by which to study disparities in spatial access to surgical care. Spatial access to healthcare is defined as the ease of individuals to access healthcare services based on the locations of populations and healthcare providers, and involves both travel cost (e.g. distance, time) and supply-demand relationships (e.g. population demand, hospital capacity, resources). EGS conditions, with their time-sensitive and resource-intensive nature may therefore be particularly sensitive to disparities in spatial access to care. While commonly-used measures of travel cost (i.e. travel distance, time) or regional availability are straightforward and easy to understand, they do not capture the effect of supply and demand. Models that reflect both aspects are needed to understand spatial access to surgical care, but have not been widely studied in this field.

To address this gap, we sought to evaluate the utility of a gravity-based spatial access model in measuring spatial access to EGS. Using California as a sample state due to its large size and diverse population, we compared the Spatial Access Ratio, a gravity-based model that accounts for travel cost and supply-demand relationships, to a single travel cost model in first their ability to discern patterns of spatial access to EGS-capable hospitals, and secondly, their ability to meaningfully identify disparities in spatial access. We hypothesized that the spatial access model would show a greater ability to identify disparities for the population as a whole, as well as for rural and vulnerable groups.

## METHODS

### Geospatial Data

We first constructed a GIS platform for EGS-capable hospitals in California. Geospatial data for the state of California including census units, zip code, city boundaries and transportation networks was obtained from the Census Topologically Integrated Geographic Encoding and Referencing file and the StreetMap North America network data set from the Environmental Systems Research Institute.^7,8^

### Hospital Identification

We used the American Hospital Association 2015 Annual Survey Database to obtain information on precise geographic location, clinical resources and measures of utilization. We identified non-federal general hospitals that performed a minimum of 10 inpatient surgeries per year. Limited service hospitals and those with missing or unclear inpatient surgical capacity were excluded. We then excluded any hospital without an emergency department, or general surgery service. Presence of these services were verified by manual review of hospital websites, or email communication where appropriate. Hospitals were classified as having “advanced” resources if they had at least 5 general ICU beds, at least one full time intensivist, the presence of a CT scanner, ultrasound and advanced gastroenterology services (identified by Endoscopic Retrograde Cholangiopancreatographcy (ERCP)). These resources were selected for their relevance to management of EGS diseases, and patients with complex or severe illness. This information was obtained from the AHA database, and if missing or unavailable, review of hospital websites or email communication. Annual inpatient surgical volume was used as an indicator of surgical capacity for calculation of the Spatial Access Ratio (SPAR).

### Socio-demographic Data

Socio-demographic characteristics examined included population size (overall population and population by race/ethnicity), poverty rate, health insurance coverage rate, and rural/urban status at the census block group (CBG) level. CBGs are the smallest geographical unit for which the census bureau publishes sample data. Population, poverty, and health insurance data were collected from the American Community Survey 2003–2007 five-year average dataset through the IPUMS National Historical Geographic Information System^9^. For race/ethnicity, we categorized the population into five mutually exclusive groups: non-Hispanic White (NHW), non-Hispanic Blacks (NHB), Hispanic (H), Asian (A) and Native American/American Indian (NA). The health insurance coverage rate was calculated as the percentage of those age 19–64 who have at least one type of health insurance (e.g., employer based, privately purchased, Medicaid, Tricare or military-based, and Veterans Administration-based).

Rural/urban status of CBGs were derived from the census tract (i.e., a higher level of census units aggregated from CBGs) level 2010 Rural Urban Commuting Area (RUCA) codes^10^. RUCA categorizes census tracts into four groups (i.e., metropolitan, micropolitan, small rural, and isolated rural) based on census rurality definitions and working commuting data. For each CBG, the rural/urban status was determined by that of the census tract it falls in. Disparities spatial access were assessed using both Shortest Travel Time Model and the Spatial Access Ratio, described below.

### Shortest Travel Time Model

Travel time between a population site and an EGS site was calculated using the ArcMap software (version 10.5)^11^. Each CBG is considered as a population site and generally contain between 600 and 3,000 people. A population-weighted centroid based on lower level (i.e., census block) population was generated for each CBG to represent the population site’s location. Then, the travel time from a population site to an EGS hospital was calculated by the Network Analyst in ArcMap based on street network data and speed limit information^12^. As we did not have detailed address information for each individual, we made the *a priori* assumption that all individuals within a CBG would have the same travel time to the nearest EGS hospital. After calculating travel times from a population site to all EGS hospitals, the shortest travel times to any EGS hospitals and advanced EGS hospitals was selected. Categories of travel time were set at 10 minute intervals up to 30 minutes, then from 30.1–60 minutes, 60.1–120 minutes and > 120 minutes. Optimal travel was defined *a priori* as < 20 minutes, while prolonged travel time was defined as > 60 minutes.

### Spatial Access Ratio Model

Gravity models of spatial access account for complicated interactions among healthcare supply, population demand, and travel cost between population locations and healthcare sites^12–16^. Gravity-based spatial access models estimate spatial access to medical services based on the law of gravitation^17^. Specifically, gravity models assume a population site’s spatial access to a medical site decreases with the increase of travel distance to that medical site. A distance impedance function, *f(d)*, is generally used to model the influence of travel distance *d* on the spatial access. One of the most commonly used and widely validated gravity models is the enhanced 2-step floating catchment area (E2SFCA) method^15^. The details of the E2SFCA are provided in supplementary material (Suppl. 1). In brief, this two-step processes first calculates the supply-demand ratio of each medical site, and uses this to calculate a Spatial Access Index (SPAI) for each population site. This method implements the idea of gravity assumption, as a shorter distance denotes a higher population demand and better spatial access for a population site.

To calculate spatial access to EGS services in California, we used the following measures in our model. The population size of each CBG was used to approximate the population demand, while annual volume of inpatient surgical operations was used as a proxy measure of surgical capacity for each hospital, as explained above. To account for the ‘edge effect’ (i.e., individuals living in boundary areas could go to the neighboring state for medical care) in spatial access modeling, a 50-mile buffer zone was extended from the boundary of California. Both California and the buffer zone was included in the spatial access calculation, however only California CBGs are presented in our results. To minimize the influence of the distance impedance problem (i.e., the selection of the impedance parameter β could influence the spatial access results), we used a weighted spatial access index, Spatial Access Ratio (SPAR)^16^. The SPAR for a population site is calculated as the ratio between that population site’s SPAI and the average of SPAI among all population sites in the study area. The higher the SPAR, the better the spatial access. And SPAR values greater than one mean better-than-state-average spatial access, and vice versa. SPAR has been proved effective in overcoming the distance impedance problem in multiple studies and has been used to explore spatial access to a variety of healthcare services^12,16,18,19^. Categories of SPAR were identified using Jenk’s natural breaks classification method, which seeks to reduce the variance within classes, while maximizing the variance between classes. Reduced access was defined as the lowest 3 categories of spatial access out of pre-defined six classes. For the analysis of all EGS hospitals, this corresponded to an SPAR of < 0.75, and for advanced hospitals, an SPAR or < 0.81.

The study was reviewed by the University of Utah Institutional Review Board and received exempt status.

## RESULTS

### EGS Capable Hospitals

The process identified a total of 283 hospitals with EGS capability, among which 142 were with advanced EGS capability. Figure 1 shows the spatial distribution of all EGS hospitals and advanced EGS hospitals in California. Basic characteristics of hospitals are shown in Table 1. Advanced EGS hospitals were primarily located in the urban centers of San Francisco, Los Angeles and Sacramento. Only 8 (5.6%) were located in non-metropolitan areas.

**Figure 1.**
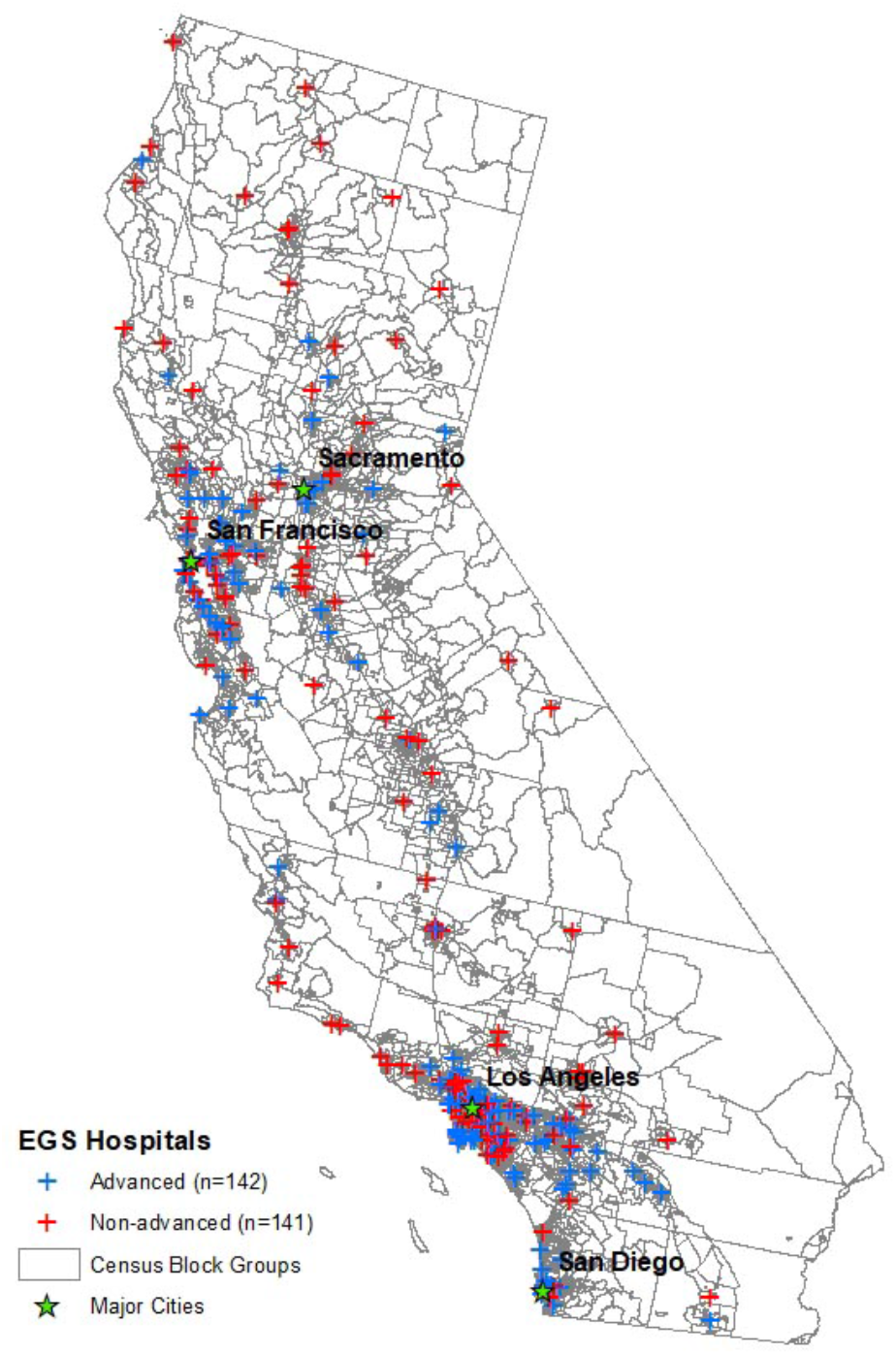
EGS hospitals in California, 2015

**Table 1.** Hospital characteristics of Basic and Advanced EGS hospitals*.

|  | **Basic** | **Advanced** | **p-value** |
| --- | --- | --- | --- |
| **Total bed number** | 139 [90-223] | 381 [249-381] | <0.01 |
| **Medical/Surgical ICU Beds** | 5 [4-10] | 18.5 [12-30] | <0.01 |
| **Emergency Department visits** | 21,194 [12,249-42,886] | 51,233.5 [34,299 – 74,985] | <0.01 |
| **Number of Operating Rooms** | 4 [3-7] | 11 [7-18] | <0.01 |
| **Number of annual inpatient surgeries** | 1304 [735 – 1970] | 2897.5 [ 1677 – 5166] | <0.01 |
| **Teaching hospital (n=184)** | 36 (25.5%) | 61 (43.0%) | <0.01 |

\*All values shown are Median [Interquartile Range] unless otherwise noted

### Spatial Access using Shortest Travel Time

Figure 2 displays the geographic patterns of shortest travel time to all EGS-capable hospitals (Figure 2a) and advanced EGS hospitals (Figure 2b). 36.98 million (94.8%) of 39 million people in California live within an optimal radius of 20 minutes driving time to an EGS-capable hospital and are primarily located in metropolitan areas or major cities. In contrast, only 0.17 million persons (0.44%) live outside a 60-minute radius of any EGS-capable hospital. These regions are predominantly located in the northern and eastern regions of the state. Travel time to advanced EGS hospitals (Figure 2b) follows a similar geographic pattern as EGS hospitals overall, however there is an increase in the proportion of people with limited access. Fewer residents were within a 20-minute radius of advanced-resource center (33.49 million, 85.8%), while over one million California residents, primarily live outside a 60-minute radius of the nearest advanced-resource EGS hospital (1.05 million, 2.69%).

**Figure 2.**
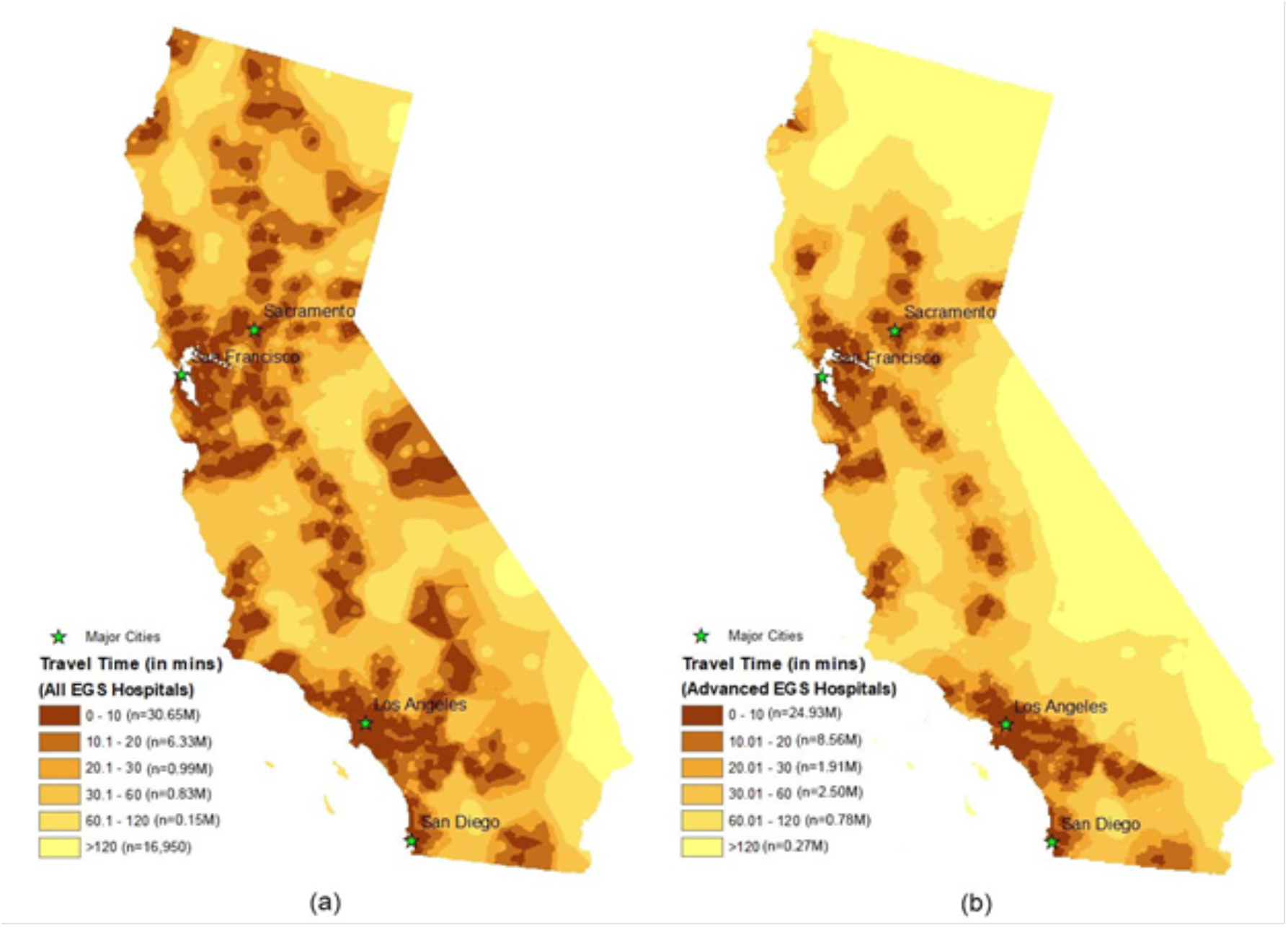
Shortest travel time to all EGS hospitals (a) and advanced EGS hospitals (b) in California, 2015 (Note: travel times were first calculated for census block group centroids and then interpolated to the entire state using an inverse distance weighting method)

### Spatial Access using Spatial Accessibility Ratio

Spatial access measured by SPAR to any EGS-capable hospital, and advanced-resource hospitals is shown in Figure 3a and b, respectively. While the overall geographic pattern of spatial access is similar to that of travel time, SPAR revealed a greater population spread across all levels of access. Using this metric 2.99 million (7.67%) people were in the category of greatest spatial access, while 11.5 million (29.5%) of people had reduced spatial access (corresponding to SPAR < 0.75) to any EGS hospital, with 1.39 million (3.56%) in the lowest category of access (SPAR < 0.2). When considering access to advanced-resource hospitals, we see once again a reduction in spatial access overall, with 13.93 million (35.7%) with reduced spatial access (corresponding to SPAR< 0.81) and 3.47 million (8.89%) in the lowest category (SPAR< 0.15).

**Figure 3.**
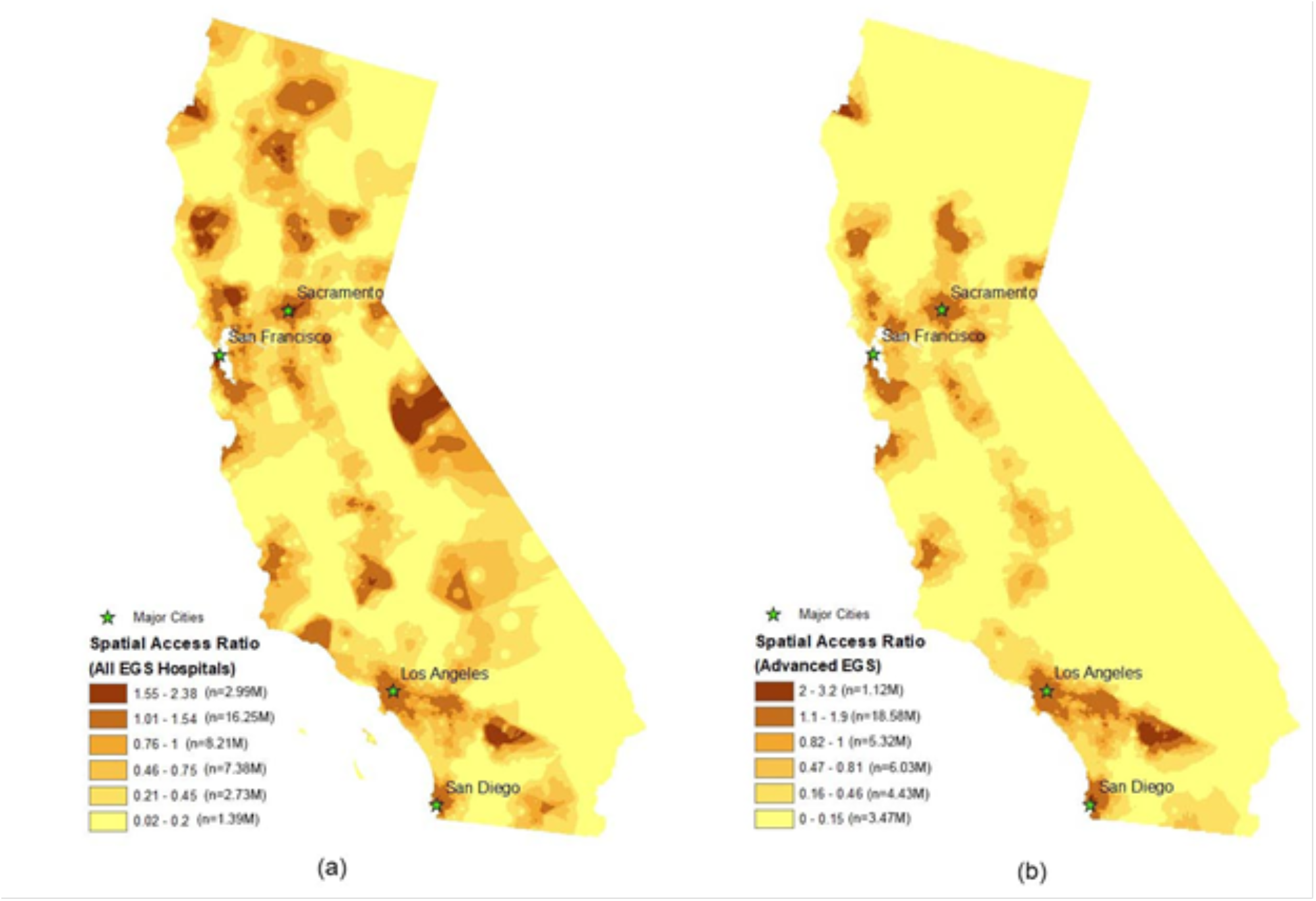
Spatial Access Ratio to all EGS hospitals (a) and advanced EGS hospitals in California, 2015 (Note: SPAR values were first calculated for census block group centroids and then interpolated to the entire state using an inverse distance weighting method; SPAR values were categorized using the Jenk’s Natural Breaks method with minor revisions)

### Patterns of Disparities in Spatial Access

Examining disparities in spatial access to EGS hospitals by travel time and SPAR show similar patterns when examining differences by race/ethnicity, socioeconomic status, health insurance coverage and rurality (Table 2). Hispanic and Native American/American Indian persons show slight, but non-significant reductions in spatial access to any EGS-capable hospital compared to the overall population. Socioeconomic status and health insurance coverage show similar patterns, with once again no significant difference compared to the overall population. When considering rurality, people living outside metropolitan regions (i.e. micropolitan, small rural or isolated rural residents) had significant reductions in spatial access. Residents in isolated rural areas had a median SPAR of 0.23 [IQR 0.02–0.32] indicating spatial access only 23% of the state average, and travelled a median of 42.1 minutes [IQR 20.6–57.8] to reach any EGS-capable hospital (vs 6.85 minutes (IQR 3.52, 8.56) for metropolitan residents. Patterns of spatial access to advanced EGS services follows a similar trend, with more marked reductions in access for non-metropolitan populations (Table 2).

**Table 2.** Spatial access to EGS services in California by race/ethnicity, socioeconomic status, health insurance coverage, and rurality.

|  | Travel Time in Minutes |  | Spatial Access Ratio (SPAR)* |  |
| --- | --- | --- | --- | --- |
|  | Any EGS Hospitals<br>(median (IQR)) | Advanced EGS<br>Hospitals<br>(median, (IQR)) | Any EGS<br>Hospitals<br>(median (IQR)) | Advanced EGS<br>Hospitals<br>(median, (IQR)) |
| **By Race/Ethnicity** |  |  |  |  |
| Non-Hispanic White (n=14,760,407) | 9.02 (3.86, 10.39) | 21.04 (4.98, 15.33) | 0.93 (0.61, 1.24) | 0.91 (0.46, 1.31) |
| Non-Hispanic Black (n=2,160,157) | 6.82 (3.61, 7.84) | 18.82 (4.74, 11.30) | 1.06 (0.80, 1.34) | 1.08 (0.73, 1.47) |
| Hispanic (n=15,101,689) | 7.39 (3.40, 8.32) | 13.50 (4.80, 12.31) | 0.99 (0.70, 1.28) | 1.00 (0.62, 1.39) |
| Asian (n=5,425,238) | 6.26 (3.40, 8.01) | 18.82 (4.53, 10.86) | 1.07 (0.81, 1.31) | 1.10 (0.79, 1.46) |
| Native American/American Indian (n=137,710) | 14.92 (4.07, 15.24) | 35.18 (5.83, 39.18) | 0.83 (0.47, 1.20) | 0.73 (0.08, 1.19) |
| **By SES (Poverty Rate)** |  |  |  |  |
| Q1 (High SES) (n=9,030,489) | 8.00 (4.16, 9.97) | 19.03 (5.27, 13.06) | 0.92 (0.64, 1.19) | 0.93 (0.60, 1.26) |
| Q2 (n=10,021,864) | 8.08 (3.81, 9.75) | 16.17 (5.10, 13.13) | 0.95 (0.67, 1.23) | 0.96 (0.61, 1.33) |
| Q3 (n=10,052,938) | 7.76 (3.38, 8.53) | 19.02 (4.59, 12.82) | 1.00 (0.71, 1.29) | 1.01 (0.62, 1.42) |
| Q4 (Low SES) (n=9,850,484) | 7.61 (3.14, 7.72) | 16.97 (4.43, 12.74) | 1.04 (0.71, 1.37) | 1.04 (0.54, 1.49) |
| **By Health Insurance Coverage** |  |  |  |  |
| Q1 (High Coverage Rate) (n=8,960,075) | 8.29 (4.00, 10.10) | 18.42 (5.09, 13.60) | 0.95 (0.65, 1.22) | 0.94 (0.58, 1.32) |
| Q2 (n=10,039,293) | 7.97 (3.83, 9.55) | 23.17 (4.94, 13.46) | 0.96 (0.66, 1.23) | 0.94 (0.56, 1.33) |
| Q3 (n=9,991,152) | 7.75 (3.50, 8.60) | 16.73 (4.73, 12.94) | 0.97 (0.69, 1.27) | 0.97 (0.59, 1.38) |
| Q4 (Low Coverage Rate) (n=9,965,255) | 7.47 (3.20, 7.54) | 12.79 (4.61, 11.48) | 1.04 (0.76, 1.35) | 1.07 (0.70, 1.46) |
| **By Rurality** |  |  |  |  |
| Metropolitan (n=36,738,421) | 6.85 (3.52, 8.56) | 15.61 (4.70, 11.77) | 1.01 (0.72, 1.29) | 1.02 (0.68, 1.42) |
| Micropolitan (n=1,450,640) | 18.58 (4.59, 26.68) | 41.81 (20.18, 58.09) | 0.56 (0.13, 0.81) | 0.39 (0.01, 0.52) |
| Small Rural (n=369,726) | 29.17 (12.60, 37.91) | 81.53 (34.16, 111.03) | 0.52 (0.06, 0.89) | 0.15 (0.00, 0.07) |
| Isolated Rural (n=396,988) | 42.11 (20.64, 57.76) | 70.59 (35.40, 84.05) | 0.23 (0.02, 0.32) | 0.11 (0.00, 0.08) |

\*(note: a larger spatial access ratio (SPAR>1) represents better spatial access, and vice versa)

### DISCUSSION

Emergency general surgery encompasses a wide spectrum of diseases that are often both time-sensitive and resource-intensive. Models of spatial access that incorporate not only travel time, but also population demand and hospital capacity are particularly relevant for this surgical specialty. In this study comparing the use of the gravity-based spatial access ratio to travel time alone to assess spatial access to EGS services in California, we found the spatial access ratio showed similar overall patterns as travel time, but greater granularity in describing disparities in spatial access. Our analysis also revealed significant disparities in spatial access to both basic and advanced EGS services in California, with rural residents and Native Americans experiencing the greatest barriers.

Prior studies of spatial access to EGS services in the United States have also found large variations across geographic regions, and revealed significant disparities in access for racial/ethnic minority and socioeconomically vulnerable populations. Diaz *et al* in their study of geospatial access to inpatient surgery found an 82% increase in the number of people living greater than 60 minutes from a surgical hospital over the decade from 2005 to 2015^20^. Over this time period, there was a 12% decrease in number of surgical hospitals in the United States, and by 2015, 6.2 million people (3.9%) lived outside a 60 minute radius from any surgical hospital and 25.9 million (8.4%) from a major surgical hospital. Uribe-Lietz *et al* found that approximately 12% of Californians live in counties with limited access to surgical and anesthesia providers.^21^ An analysis of county-level EGS availability by Khubchandani *et al* demonstrated that counties with higher proportions of rural, black, Hispanic, poor and uninsured patients had significantly higher odds of not having an EGS hospital.^6^ Both studies found great variation in access patterns on a national level – particularly between the Intermountain and Midwest versus the coastal regions. Together, these studies strongly indicate limitations in access to EGS services that affect millions of U.S residents, with a differential effect on vulnerable populations. However, purely cost or regional-availability measures are limited in their ability to account for health-seeking behaviors, such as the common event of crossing county borders, or complex interactions with resource supply and demand.

To our knowledge, this is the first study to examine spatial access to surgical services using a gravity-based model, extending our knowledge from previous studies by incorporating measures of hospital capacity and population demand. Our results for spatial access to EGS in California align with the travel time-based assessment of Diaz *et al*., with 3.56% and 8.89% of individuals in the lowest category of access for all EGS and advanced-resource hospitals respectively using SPAR, but give greater granularity with regards to levels of access.^20^ SPAR offers an improved distribution of the population across the range of the metric, with substantial proportions of residents in each category, as compared to travel time, where a large degree of clustering occurs at the extremes of the scale. Cost-based measures such as travel time or distance are certainly still of value – they are simple to measure and easily understood – but may underestimate disparities in access by not accounting for capacity of the health care system or population needs.

Consideration of hospital resources and capacity in spatial access metrics is particularly important for surgical specialties such as EGS, where severely ill patients may require operating room and ICU bed availability that can be significantly affected by population demand. SPAR has been previously used to assess spatial access to medical oncologists for colorectal and breast cancer patients, with limited spatial access associated with decreased survival for rural patients.^16,22–24^ Our study results suggest that this metric can successfully be extended to surgical populations by incorporating simple measures of inpatient surgical capacity and advanced-level resources. Similar to previous studies, using the SPAR in our analysis highlights the vast disparities in spatial access to EGS care for rural residents and Native Americans.

One of the central findings in our analysis of disparities is the magnitude of limitations to EGS care for rural residents. Compared to residents of metropolitan areas, those living in small rural communities had significantly reduced spatial access to EGS care. In addition to longer travel times to reach surgical care, the overall spatial access for small rural community residents ranged from a SPAR of 52% of the state average for any EGS hospital, and only 15% for advanced level care. The rate of rural hospital closures in the United States has been increasing over the past decade, with 128 rural hospitals with inpatient capabilities closed since 2010.^25^ Such closures reduce access to not only inpatient care and surgical services, but also significantly impact pre-hospital care by increasing Emergency Medical Services (EMS) activation and transport times. Increases in EMS response times have been tied to worse clinical outcomes, including increased mortality.^26,27^ Kaufman *et al* estimate that over 1.7 million Americans were affected by rural hospital closures from 2010–2014, a number that continues to increase on an annual basis.^28^ The marked disparity we observed in spatial access to emergency surgical care is particularly worrisome for rural residents, placing them at risk for worse clinical outcomes due to delayed intervention alone.

Our results must be interpreted in the context of several limitations. First, we relied on the AHA Annual Hospital Survey for identification of eligible hospitals, and while the AHA contains data on over 85% of all hospitals in the United States, we may have missed hospitals that do not report to this organization. Secondly, while we attempted to verify hospital EGS capabilities using multiple sources of information, misclassification of hospitals – either as “EGS-capable” or “advanced” is still possible. We intentionally used a broad definition of advanced resources to avoid misclassification due to high rates of missing data for hospital resources on the AHA survey. Studies using similar methods have overall found a low rate of misclassification of EGS-capable hospitals and should not significantly affect our ability to compare the travel cost and gravity-based models.^6^ With respect to construction of the SPAR metric, we did not have EGS specific case-volume of EGS surgeons for each hospital, and therefore used overall inpatient surgical case volume as a proxy measure for hospital capacity. This metric also represents only potential spatial access to care, and does not account for individual patient characteristics such as insurance status. Insurance coverage is an important consideration that may influence patient decisions on where to seek care, but under the Emergency Medical Treatment and Labor Act should not restrict them from receiving emergency care at any hospital. As efforts to develop a national EGS registry progress, more reliable data will allow for further refinement of SPAR for this field. Finally, the SPAR is a ratio, and as such, the values we present are relative to the California state mean. Without clear benchmarks on what constitutes “adequate” spatial access to care, it is therefore difficult to delineate exactly what level of SPAR demands improvement. Nevertheless, this ratio does work well for identifying the magnitude of disparities amongst vulnerable populations in a manner that is not easily communicated by travel time or distance alone.

There is considerable potential for SPAR to influence the equitable allocation of surgical resources. Modeling the effects of adding EGS-capable hospitals to low-SPAR regions, and those of adding specific resources (e.g. operating rooms, critical care capacity), will allow for estimations of how these efforts will affect the overall population, and reduce known disparities in access to care.^29–32^ In addition to evaluating how potential spatial access compares to realized access to care, further work is needed to examine the contribution of spatial access to clinical outcomes, and influences of sociodemographic factors on this relationship. This understanding will be critical to developing coordinated care strategies that optimize outcomes for all EGS patients, including those who experience the greatest challenges reaching the hospital in the first place.

### CONCLUSIONS

Models of spatial access that incorporate measures of hospital capacity and population demand in addition to travel cost are particularly relevant to surgical fields such as EGS that require physical resources to treat patients. While travel time and SPAR showed similar overall patterns of spatial access to EGS-capable hospitals in California, SPAR showed greater differentiation of access across the state, and may be particularly useful in evaluating disparities and directing allocation of resources. Nearly one-third of California residents has limited or poor access to EGS hospital, with the greatest disparities noted for rural residents.

## Data Availability

The authors confirm that the data supporting the findings of this study are available within the article and its supplementary materials.

## FUNDING/ACKNOLWEDGEMENTS

Dr McCrum was supported by funding from the American Association for the Surgery of Trauma (AAST) Research and Education Scholarship and the National Institute of Health (Grant #R21MD012657).

